# AMU: Using mRNA Embedding in Self-Attention Network to Predict Melanoma Immune Checkpoint Inhibitor Response

**DOI:** 10.1101/2022.10.01.22280593

**Authors:** Yi Yin, Qing Wu, Ziming Wang, Yu Kang, Xianhe Xie

**Affiliations:** Department of Medical Oncology, Clinical Oncology School of Fujian Medical University, Fujian Cancer Hospital; Fuzhou, 350014, China; Department of Oncology, Molecular Oncology Research Institute, Fujian Key Laboratory of Precision Medicine for Cancer, The First Affiliated Hospital of Fujian Medical University; Fuzhou, 350005, China; School of Information and Communication Engineering, Beijing University of Posts and Telecommunications, Beijing, 100876, China

**Keywords:** deep learning, transformer, self-attention, immunotherapy, representation learning

## Abstract

**Background:** To precisely predict drug response and avoid unnecessary treatment have been urgent needs to be resolved in the age of melanoma immunotherapy. Deep learning model is a powerful instrument to predict drug response. Simultaneously extracting the function and expression data characteristics of mRNA may help to improve the prediction performance of the model.

**Methods:** We designed a deep learning model named AMU with self-attention structure which were fed with the mRNA expression values for predicting melanoma immune checkpoint inhibitor clinical responses.

**Results:** Comparing with SVM, Random Forest, AdaBoost, XGBoost and the classic convolutional network, AMU showed the preferred performance with the AUC of 0.941 and mAP of 0.960 in validation dataset and AUC of 0.672, mAP of 0.800 in testing dataset, respectively. In model interpretation work, TNF-TNFRSF1A pathway were indicated as a key pathway to influence melanoma immunotherapy responses. Further, gene features extracted from embedding layer and calculated by t-SNE algorithm, showed a local similarity with Functional Protein Association Network (STRING, https://cn.string-db.org/), AMU could predict gene functions and interactions simultaneously.

**Conclusions:** Deep learning model built with self-attention structure has strong power to process mRNA expression data and gene vector representation is a promising work in biomedical field.

**What is already known on this topic:** The types of biomarkers for immunotherapy are very complex and transcriptomics biomarker research is one part of it, but currently it is lack of generally acknowledged results with practical value. Combining deep learning models with transcriptomics biomarker markers can help us to predict drug sensitivity. However, the powerful capabilities of deep learning models have not been fully exploited and utilized.

**What this study adds:** The expression of 160 genes could well predict the efficacy of immunotherapy, even if the tissue samples were after drug administration, and through model training, we could also extract the interactions and connections between genes. The deep learning model could not only do prediction, but were also promising in performing gene vector representation learning.

**How this study might affect research, practice or policy:** Our research is not only to provide a model with high predictive value, but also to extract gene interaction relations during model training, which is very enlightening for gene vector representation learning. The research of gene vector representation learning can promote the prediction accuracy of deep learning models in various biomedical fields because it can become the common upstream of many biomedical tasks.

## BACKGROUND

The publication of Alexnet in 2012 brought neural network back to researchers’ attention,[1]. After ten years of development, Deep learning (DL), a computer science and technology with the neural network as the core, has become one of the most active scientific research fields and the primary technology of artificial intelligence. It takes a big step forward in the development of artificial intelligence (AI), and promotes great changes and progress in the fields of industry, agriculture, commerce, economic finance, and medical area etc. Deep learning-centered artificial intelligence has become a key technology in the new industrial revolution.

DL has developed rapidly in image recognition and image segmentation, and has already been mature and widely used in industry. In the field of Natural Language Processing (NLP), the self-attention mechanism was proposed by Google in its famous paper “ Attention is All Your Need” in 2017, which is conducive to integrate the internal association between the input long sequence data and improve the predictive accuracy of the downstream tasks such as automatic speech recognition, machine translation etc. Subsequently, the transformer network with self-attention mechanism as the core architecture was widely proved to be superior and quickly became one of the acknowledged optimal basic networks. Then the transformer was transplanted to computer vision (CV) field and models such as ViT and Swin Transformer were proposed,[2, 3], which greatly improved the prediction accuracy of CV tasks. In the medical field, CV model or NLP model with transformer is also widely used,[4], AI assisted pathological/ imaging diagnosis and medical data extraction are under accelerated development,[5]. Meanwhile, in the field of scientific research, Graph neural network (GNN) models is often applied to drug sensitivity prediction and molecule affinity prediction,[3, 6]. However, AlphaFold2 with self-attention mechanism successfully predicted protein tertiary structure based on protein primary structure information last year,[7], which is believed that it can greatly improve the efficiency of protein function studies. For gene multi-omics data, most of current studies feed the data into the Convolutional Neural Network (CNN) in the form of one hot coding or one-dimensional vector,[3, 8, 9], which seems lags behind other areas in DL domain. In this study, considering the interaction and connection among genes, we tried to use transformer encoder structure with self-attention mechanism to manage gene expression data, which achieved good prediction results and suggested the feasibility of self-attention mechanism for gene vector representation.

We named our DL model AMU, which meant Attention mechanism Model for predicting melanoma iMMUnotherapy checkpoint inhibitor (ICI) response. In recent years, malignancy immunotherapy had made great progress and significantly improved patients’ overall survival, especially for melanoma, immunotherapy had already acquitted as the standard treatment in the advanced disease,[10, 11]. However, the clinical tumor response to immunotherapy is not satisfied, and the objective response rate (ORR), which is the standard assessment criteria for evaluating anti-tumor drug activation, is around 30%,[12], in some other tumors, the ORR is even lower, around 10%-20%,[13, 14]. So how to precisely identify which group of patients can beneficial from immunotherapy has caused much attention,[15-17]. Currently approved immunotherapy treatment includes PD-1/PD-L1 inhibitor and CTLA-4 inhibitor, the biomarkers for these drugs are usually PD-L1 expression level, tumor mutation burden (TMB) and MSI-H/dMMR status, but these biomarkers have low prediction accuracies and often contradict with each other,[18-20]. The search for more precise methods has not stopped, we considered drug response is related to complex biological pathways and conducted this study using muti-gene mRNA expression values to predict ICI response.

We summarized our contributions follows:

1. In model building and developing level:
  - We provided reliable evidence to reveal the superiority of our model AMU achieving excellent performance in both validation dataset and independent testing dataset for melanoma ICI response prediction, highlighting the strong predictive power and generalization ability of our model.
  - We proved that the self-attention mechanism could work in 1-D vector data, even if the input data is not spatial positional type image or sequence sensitive type natural language.
  - We discovered the embedding architecture could be used for representation learning of gene feature and combining mRNA expression quantitative information, the interactions of learned representation vector had local consistence with the widely accepted Functional Protein Association Network (STRING, https://cn.string-db.org/),[21] which proved the embedding architecture is suitable and promising for gene vector representation. Self-attention mechanism was superior and benefit for digitating data inner correlation. The interpretation of embedding layer made the DL network becoming more convincing, which was especially important in biomedical area.
2. In biological level:
  - According to model interpretation work, we put forward an assumption that the TNF-TNFRSF1A pathway might be a key pathway to decide melanoma ICI response.
  - CD80 and CCR3 expression may related to both survival and ICI response for melanoma.

## METHODS

We collected the open data to build our datasets, AMU performance was evaluated in validation and testing datasets comparing with other five machine learning models, after the model was trained, we conducted the interpretation work to explore the importance and interactions of gene functions. We used DL framework PaddlePaddle 2.3.0 to build AMU and Paddle AI Studio (https://aistudio.baidu.com/aistudio/index) to train model online, figures were drawn by matplotlib package.

### Overview of AMU framework

AMU is constructed by a transformer encoder followed with a convolutional network for an ICI clinical response binary classification task. The input data are 160 normalized mRNA expression values. As the same as other classification models, the output of AMU is a pair of probability values, which denotes non-response and response probability. The transformer encoder structure is classic as that in NLP, which will be described in detail in the following part. In convolutional network, we used ‘Convolution-Dropout - Batch Normalization - ReLU activation function - Adaptive Maximum Pool’ strategies. We used Adam algorithm as the optimizer for back-propagation process and two-step decay of learning rate for training. AMU takes the SoftMax activation function for the end of the net and cross-entropy as loss function. A total of 83,462 parameters are trainable in AMU. Details see Supplement (Table S1).

### mRNA embedding and transformer encoder layer

We set a 20-D gene embedding for gene feature learning, and the initialized embedding input is integer “1” to “160”, then we multiply embedded values with mRNA expression values in order to add expression information to every embedding, this method was inspired by NLP process in which word position information is added to the embedding layer. We consider that gene features can be learned in the end-to-end training process just like words can be representation learned in large text corpus. However, gene is not a sequence data so that the position information is not necessary and expression information should be instead. Genes have interaction and association with each other, so self-attention mechanism will be work.

In the process of transplanting transformer encoder layer, no structure needs to be changed, which includes Layer normalization, Dropout, Muti-head attention, and Multilayer Perceptron (MLP). In model training experiment, we used eight Muti-head attentions and repeated transformer encoder layer eight times to avoid underfitting.

### Building Dataset

As show in Table 5, all the cases fed into models are collected from published data, including three independent datasets GSE78220, GSE91061, GSE165278 from GEO Datasets and one dataset from the paper of Liu (PMID:31792460),[22-26]. We collected total 206 patients diagnosed with advanced melanoma treated with immunotherapy checkpoint inhibitor, including Nivolumab, Pembrolizumab and Ipilimumab. The whole-transcriptome sequencing (RNA-seq) conducted on pretreatment tumor tissues.

The testing dataset was built by 58 post-treatment tissue samples from GSE91061. Clinical information is not available in the most datasets and the patients’ characteristics cannot be described. Datasets detail are shown in **Table 1**.

**Table 1.** Summary of datasets.

| Dataset ID | Patient<br>count | Drug applied | Biopsies type | Sample<br>acquisition | mRNA-seq<br>platform |
| --- | --- | --- | --- | --- | --- |
| **Training/Validation dataset (n = 206)** |  |  |  |  |  |
| GSE78220 | 27 | Pembrolizumab | Melanoma tissue | pre-anti-PD-1<br>therapy | Illumina HiSeq<br>2000 |
| GSE91061 | 51 | Nivolumab | Melanoma tissue | pre-anti-PD-1<br>therapy | Illumina<br>Genome<br>Analyzer |
| GSE165278 | 7 | Ipilimumab | Melanoma tissue | pre-anti-CTLA-4<br>therapy | Illumina HiSeq<br>2500 |
| Liu | 121 | Nivolumab/<br>Pembrolizumab | Melanoma tissue | pre-anti-PD-1<br>therapy | Illumina HiSeq<br>2000/ 2500 |
| **Testing dataset (n = 58)** |  |  |  |  |  |
| GSE91061 | 58 | Nivolumab | Melanoma tissue | post-anti-PD-1<br>therapy | Illumina<br>Genome<br>Analyzer |

According to previous studies, 169 genes have been described potential associated with melanoma, inflammation, immunity, the PD-L1/CTLA4 pathways and ICI response,[27, 28, 29]. Finally, 160 genes were overlapped in four datasets and selected. For response digital representation, we took records with “complete response (CR)” and “partial response (PR)” as response (classed as numeric 1), “stable disease (SD)” and “progression disease (PD)” as non-response (classed as numeric 0). Supplement (Data file S1) lists the full 160 gene names.

We calculated the TPM-normalized expression values by the raw data provided by authors. And continued to normalize the TPM values to constituent ratios, then we selected the 160 values from their original datasets and convert to constituent ratios again. After this step, all the values from different datasets are represent mRNA relative expression quantity and are comparable. At last, we logarithm them.

The last, we applied up sample strategy for data enhancement, positive samples were 1:1 duplicated. A total of 280 samples were in our training/validation dataset including 148 positive samples and 132 negative samples. We randomly divided the training/ validation dataset into the training (224) and validation (56) sets, which corresponded to 80% and 20% of the total instances, respectively. In order to get reliable model performances, we randomly split training and validation data five times, which will be mentioned as “5-fold cross validation” in the following part.

### Competing methods

We chose four machine learning models and designed one simple CNN as competing methods. All the competing models input data are 160-D vector of mRNA expression values we have been described.

- SVM (Support Vector Machine) is a first-class classification machine learning model. We employed the grid search strategy to find the optimal model hyperparameter. ‘kernel’ included ‘linear’, ‘poly’ and ‘rbf’, ‘C’ was in the list [1, 10, 100], ‘gamma’ was in the list [1, 0.1, 0.001].
- Random forest is a tree-based regressor model. We set the number of trees in the forest from range (2,10) and ‘n_estimators’ from arrange (10,300,10). The best hyperparameters were chosen in the comparing experiments.
- AdaBoostClassifier is a tree-based ensemble model and the best hyperparameters were chose as the same as Random Forest.
- XGBoost (eXtreme Gradient Boosting) is a scalable tree boosting system. It implements machine learning algorithms under the gradient boosting framework. One of the advantages of XGBoostClassifier is convenience for model interpretation,[30].
- CNN, we built a simple CNN to represent traditional DL model without self-attention mechanism. The model included three Conv1D layers and total 737 trainable parameters. Details see Supplement (Table S2).

### Model evaluation

In classification experiments, AUC and PR curves the two commonly used measurements were chosen as our classification metrics. To further evaluate the performance of our model, we demonstrated results under validation dataset and testing dataset respectively. We also used several common metrics in five-fold cross validations, including accuracy, precision, recall and f1 score.

### Model interpretation

We selected SVM, XGBoost and AMU model to explore model interpretation works.

For SVM and XGBoost models, we applied SHAP (SHapley Additive Explanations) which is a game theoretic approach to estimate the gene feature importance, then we used GO pathway enrichment analysis and overall survival COX analysis to describe the important gene features.

For AMU model, Shap also can identify the gene importance, but more information can analysis through mRNA embedding layer. Just be inspired by NLP word embedding, we toke mRNA embedding layer 20-D trainable parameters as gene features. We tried the cluster analysis and calculated the Euclidean distance, cosine similarity and t-distributed Stochastic Neighbor Embedding (t-SNE) among gene feature vectors to describe the gene association and interaction. The we compare the gene correlations with Functional Protein Interaction Network (STRING, https://cn.string-db.org/) to evaluate the gene feature learned from AMU.

## RESULTS

### AMU accurately predicted melanoma immunotherapy response

We identified the performance of our model on validation and testing datasets comparing with currently advanced machine learning models in five-fold cross validations. **Table 2** showed the binary classification reports of validation dataset predicted by original training data. DL models were not preferred, and SVM had the best performance according to the accuracy score (0.633) and recall score (0.633). All the models had unsatisfactory performance. XGBoost model get the highest f1-score (0.567) followed by AMU (0.55), the CNN model had the lowest f1-score of 0.45.

**Table 2.**
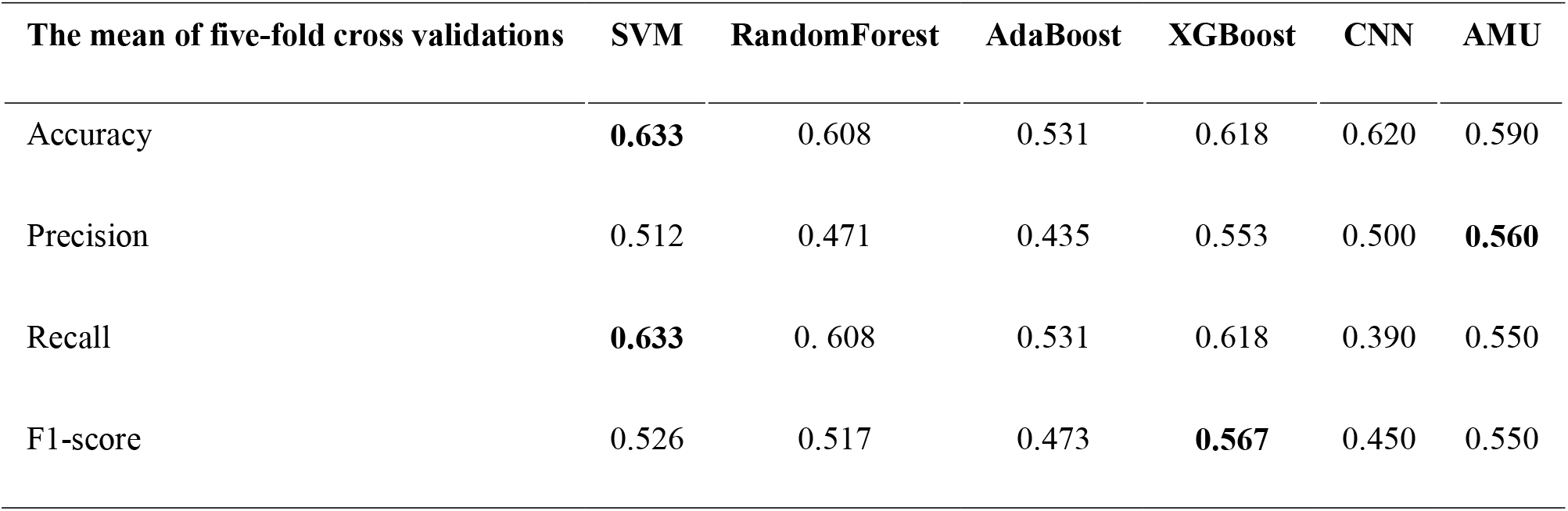
Classification reports of Amu and five comparing methods for original validation dataset.

However, after data enhancement, all the model performances were significantly improved except CNN (**Table 3**), which was hard to converge. AMU model showed the best performance with f1-score 0.93, the area under the curve (AUC) 0.941 and mean average precision (mAP) 0.960, respectively. In the testing dataset, AMU also demonstrated superior predictive perform as show in **Table 4** and achieved the highest AUC (0.672) and mAP (0.800) respectively. The receiver operating characteristic curve (ROC) and Precision-Recall (PR) curve were show in

**Table 3.** Classification reports of Amu and five comparing methods for enhanced validation dataset.

| The mean of five-fold cross validations | SVM | RandomForest | AdaBoost | XGBoost | CNN | AMU |
| --- | --- | --- | --- | --- | --- | --- |
| Accuracy | 0.884 | 0.653 | 0.821 | 0.792 | 0.544 | **0.930** |
| Precision | 0.906 | 0.707 | 0.872 | 0.854 | 0.524 | **0.930** |
| Recall | 0.892 | 0.654 | 0.821 | 0.792 | 0.534 | **0.930** |
| F1-score | 0.884 | 0.611 | 0.809 | 0.777 | 0.482 | **0.928** |

**Table 4.** Classification reports of Amu and five comparing methods for testing dataset.

|  | SVM | RandomForest | AdaBoost | XGBoost | CNN | AMU |
| --- | --- | --- | --- | --- | --- | --- |
| Accuracy | 0.76 | 0.55 | 0.64 | 0.67 | 0.71 | **0.72** |
| Precision | 0.38 | 0.45 | 0.46 | 0.44 | 0.59 | **0.61** |
| Recall | 0.50 | 0.44 | 0.47 | 0.47 | 0.59 | **0.60** |
| F1-score | 0.43 | 0.44 | 0.46 | 0.45 | 0.59 | **0.60** |

**Table 5.** Top-10 Shap value genes prioritized by SVM, XGBoost and AMU.

| Module | Top-10 Shap value genes |
| --- | --- |
| SVM | TNFRSF1A, SERPINA1, F5, NEDD4L, TLR4, SERPINE1, GYPB, NBEA, BPGM, UBE2C |
| XGBoost | FASLG, CDKN1A, TP53, CD4, CASP3, HMGB1, SLC4A1, TNF, FOS, IL5 |
| AMU | THBS1, CD86, MIF, BPGM, NRAS, TNF, IL23A, CXCL8, CD40, TNFRSF1A |

### Model interpretation

We listed the top-10 Shap value genes of SVM, XGBoost and AMU (**Table 5**)[31]. The top genes were quite different among models. The intersection of these top-10 genes was including TNF and its receptor TNFRSF1A. TNF encodes a multifunctional proinflammatory cytokine. TNFRSF1A is a member of the TNF receptor superfamily of proteins. The details of Shap values were in Supplementary (Fig S1-3, Data files S2-4).

Gene Ontology (GO) analysis was performed in top-50 genes of these models[32-34], total 112 genes were gathered, the enrichment analysis of pathways was show in **Figure 3**. The most important genes are clustered in lymphocyte proliferation pathway. Then overall survival cox analysis of these 112 genes was conducted (**Figure 4)**, 17 genes showed statistical significance, most genes showed protect effects, only 2 genes (CD 80 and CCR3) had noteworthy hazard ratios (HRs) (0.761 and 0.134 respectively). CD 80 protein was activated by the binding of CD28 or CTLA-4 and then induces T-cell proliferation and cytokine production. CCR3 protein is a receptor for C-C type chemokines.

**Figure 1.** The overview of AMU

**Figure. 2.** ROC and PR in validation dataset. (A) and (B) respectively shows ROC curve and PR curve with 6 models for validatio dataset. The PR curve shows mean average precision (mAP) of 2 classes. (C) and (D) respectively shows ROC curve and PR curve for testing dataset.

**Figure 3.** GO pathways enrichment analysis. (A): GO results of three ontologies. (B): Biological process of pathways enrichment.

**Figure 4.** Cox analysis of Top-50 Shap value genes of (SVM、XGB、AMU) models (genes of p values <0.05)

Finally, but most important, gene features learned by AMU showed biological significance. We found that mRNA embedding matrix was hard to perform a desirable cluster analysis, also, the Euclidean distance and cosine similarity algorithm both revealed the gene features distributed uniformly and no aggregation. **See details in Supplementary (Fig S4-7, Data file S 5-6)**. However, gene association and interaction calculated with t-SNE algorithm showed locally similar with STRING. Four cases were visualization in **Figure 5**. For CD4-MAPK14-PTPRC-SOCS1 subgroup, both mRNA embedding and STRING indicated inner close association, and NEDD9 relatively isolated with them. For PDE3B-ELANE-CXCL8, mRNA embedding successfully mapped the close distance. In NRAS-LAGLS3-IL10-FCGR2B-CDKN1A-HMGB1 subgroup, most links were accurately figure out with a local difference that STRING showed CDKN1A -NRAS, but mRNA embedding showed CDKN1A-FCGR2B association. Another case was in CASP1-TLR9-CXCR3-ITGAL-TXNRD1 subgroup, most links were consistent except STRING described an interaction with CXCR3-ITGAL, but mRNA embedding didn’t figure it out.

**Figure 5.** Gene interaction learned by AMU and compared with STRING (A): NRAS-LAGLS3-IL10-FCGR2B-CDKN1A-HMGB (B): CASP1-TLR9-CXCR3-ITGAL-TXNRD1 (C): PDE3B-ELANE-CXCL8 (D) CD4-MAPK14-PTPRC-SOCS1

## DISCUSSION

In industrial 4.0 age, DL has been the most advanced model algorithm. Since the Alexnet proposed in 2012, convolutional network renewed and a new wave of artificial intelligence research and applications have begun. Then transformer has been the most advanced deep learning technique and exhibited powerful performers in CV and NLP areas for its strong features extraction ability of sequential and spatial interactions of data. AMU is a model connected the transformer encoder with a convolutional network, it’s a successful trial of proving that the transformer structure is also feasible and superior for 1-D gene expression data (just like NLP) prediction task and splendid for gene feature learning. AMU also showed superior performance in testing dataset which tissue biopsies were post ICI and some of the features and data distribution had to be different from that of pre-ICI, further proved AMU learned some essential features. Previously, several works have been done in using mRNA expression and clinical data to predict melanoma ICI response. Noam Auslander etc. reported an AUC of 0.83 with their IMPRES predictor,[35]. Another algorithm proposed by Philip Friedlander etc. was validated in the validation set with AUCs of 0.62,[27]. By this cross-experiment comparison, AMU exhibited its advantage.

Further, features abstracted from embedding layer showed local similarity with laboratory results or curated databases, indicating strong gene presentation abilities of transformer encoder should be fully researched and utilized for more gene related downstream tasks. DL studies should sufficiently take advantage of the power of transformer. Moreover, model interpretation is quite important for medical studies and obviously gene embedding can facilitate this work.

Additionally, in the model interpretation part, SVM, XGBoost and AMU consistently indicated that TNF-TNFRSF1A axis possess the most important genes related to melanoma ICI response process. Previous mouse experiments published in Nature showed that anti-PD-1 and anti-CTLA-4 (NIVO+IPI) combined with TNF-α inhibitors could improve the course of colitis in a mouse model and enhance the anti-tumor effect,[36]. Phase Ib clinical trial showed the promising effect of combining Nivolumab and Ipilimumab with TNF-α inhibitor in advanced melanoma,[37]. These facts indicated machine learning models not only can applied in predictive scenarios but also can provide suggestive information for further investigation.

Our work has several limits:

1. sample size was not large enough and the representativeness of samples was inevitably impaired, meanwhile, the patients’ characteristics were not described, which will limit the extrapolation of the results. It is a common problem in medical deep learning researches because the data is limited for one specific task and unsupervised learning often works to resolve this dilemma. For DL, models have strong fitting ability, but sample diversity and distribution decide the model generalization. A larger and closer to real situation training dataset is desired for robust performance in most cases.
2. Although 160 genes were much less than previous studies imported 500-800 genes and more favorable for model interpretation, we consider that input features can be slimed more accurately because the ratio of input features and sample numbers should be controlled within a certain range for a better result according to Ben Sorscher’s paper,[38].
3. A particular point we had to indicate is that, different from other fields, input features are almost impossible to be enumerated in medical studies. Taking melanoma ICI response as example, input data can include multi-omics, clinical, pathological and imaging data etc. Our study only imported mRNA expression data, which is not complete for feature abstraction.

Looking for immunotherapy biomarkers requires multidisciplinary collaboration; our self-attention model is powerful in extraction and integration of the transcriptome information and make the drug sensitivity prediction more credible. The nature of gene is information, all kinds of advanced techniques used to process information can be tried to process gene data. In our opinion, gene representation learning work should be promising, because it can be used as a common upstream path for biological information mining and make our target tasks performed better.

## Data Availability

All data produced in the present study are available upon reasonable request to the authors
All data produced in the present work are contained in the manuscript
All data produced are available online at https://aistudio.baidu.com/aistudio/projectdetail/4298990

## Acknowledgments

The authors thank Xiaohua Zhang, Cunxi Li for helpful discussion, Tianxuan Qi for coding enlightenment, Muqun He, Jianfeng Wang and Yunjian Huang for the work incentives, and thanks to Baidu Paddle team for providing free online GPU and training courses to us.

## List of Abbreviations

AI: Artificial Intelligence
AUC: The Area Under the Curve
CV: Computer Vision
CNN: Convolutional Neural Network
DL: Deep Learning
GNN: Graph Neural Network
GO: Gene Ontology
ICI: Immune Checkpoint Inhibitor
mAP: mean Average Precision
NLP: Natural Language Processing
ORR: Objective Response Rate
PR: Precision-Recall
ROC: receiver operating characteristic curve
SVM: Support Vector Machine
SHAP: SHapley Additive Explanations
TMB: Tumor Mutation Burden
XGBoost: eXtreme Gradient Boosting

## List of Supplementary Materials

Fig. S1: SVM Top-20 Shap gene values

Fig. S2: XGBoost Top-20 Shap gene values

Fig. S1: AMU Top-20 Shap gene values

Fig. S4: Clustering analysis of AMU gene embeddings

Fig. S5: Heatmap of Euclidean distance of AMU gene embeddings

Fig. S6 Heatmap of cosine similarity of AMU gene embeddings

Fig. S7: Bar chart of cosine similarity of AMU gene embeddings

Table S1: Summary of AMU framework

Table S2: Summary of CNN framework

Data files S1: 160 gene names

Data files S2: SVM Shap values

Data files S3: XGBoost Shap values

Data files S4: AMU Shap values

Data files S5: Euclidean distance of AMU gene embeddings

Data files S6: cosine similarity of AMU gene embeddings

